# Prevalence of salivary anti-SARS-CoV-2 IgG antibodies in vaccinated children

**DOI:** 10.1101/2022.12.15.22283480

**Authors:** María Noel Badano, Alejandra Duarte, Gabriela Salamone, Florencia Sabbione, Matias Pereson, Roberto Chuit, Patricia Baré

## Abstract

Vaccination against COVID-19 has mitigated the impact of SARS-CoV-2 infection, decreasing the probability of progression to severe disease and death in vaccinated people.

Parallel to the development and administration of COVID-19 vaccines, the immune response induced by the different vaccine platforms has been investigated, mainly, in the adult population. However, since the approval of the vaccines for use in pediatric individuals was a posteriori, vaccination began later in this population. This, added to the difficulty in obtaining blood samples from pediatric individuals, has led to less knowledge about the humoral immune response following vaccination in children.

In this work, we analyzed the humoral response induced by vaccination in children through a non-invasive approach such as the measurement of specific salivary antibodies. Our results showed a high prevalence of specific salivary antibodies (81%), with the highest levels of antibodies being observed in those children who had three doses, a greater number of exposures and a shorter interval time between the last exposure to SARS-CoV-2 antigens and saliva collection. These results agree with those reported for the systemic humoral immune response in vaccinated adults, suggesting the administration of booster doses in children to maintain high antibody levels.

Therefore, determination of salivary antibodies against SARS-CoV-2 could be a non-invasive tool for disease surveillance, vaccination follow-up and to assist vaccination strategies against COVID-19.

## INTRODUCTION

Systemic humoral response following COVID-19 vaccination has been widely studied, while less attention had received salivary response. Reports showed specific salivary antibodies after vaccination with COVID-19 mRNA vaccines, which correlated with serum neutralization and indicated seroconversion.^1, 2^ However, most studies were conducted in adults with few studies about immune response following vaccination in children.^3-5^

In this study, we aimed to analyze the humoral response following vaccination in children, through a non-invasive approach like the measurement of specific salivary antibodies.

## METHODS

Since March 2022, children younger than 18 years were recruited to analyze prevalence of salivary anti-SARS-CoV-2 IgG antibodies. IgG anti-spike antibody concentrations, in binding antibody units (BAU) per mL (BAU/mL), were determined by ELISA.^6^ Statistical analyzes were performed with GraphPad Prism software. Geometric mean concentrations (GMC) of antibody levels with 95% confidence intervals (95% CI) were calculated. Antibody levels between groups were compared with Mann-Whitney test and between paired samples with Paired t test. Multiple linear regression was performed to analyze variables associated with antibody levels. Statistical significance was defined as a 2-tailed *p*< 0.05. Supplemental information includes other experimental methods. This study was approved by the Academia Nacional de Medicina Ethics Committee. Written informed consent was obtained from parents.

## RESULTS

Saliva samples were collected from 89 children (Median age: 10 (4−17); 47/89 (52%): male) including unvaccinated (n=1), with one (n=2), two (n=52) and three (n=34) vaccine doses (Table 1). Fourteen subjects had a confirmed past SARS-CoV-2 infection, 39 were household contacts and 36 were unexposed subjects. Median time between last exposure to SARS-CoV-2 antigens (through vaccination, infection or exposure) and saliva collection was 77 (21−270) days. Prevalence of SARS-CoV-2 spike-specific IgG antibodies in total samples was 81% (72/89), 73% (38/52) in children with two vaccine doses and 97% (33/34) in those with three doses. Higher antibody concentrations were observed in subjects receiving three doses compared to those receiving two (GMC: 834.0 IU/ml, 95% CI: 456.6−1523 *vs* GMC: 32.7 IU/ml, 95% CI: 17.0−63.1; *p*< 0.0001) (Figure 1A), showing highest levels those with heterologous third doses, who received two doses of Sinopharm vaccine and mRNA vaccines as third doses (GMC: 1997 IU/ml, 95% CI: 878.6−4538; *p*< 0.01). Antibody concentrations increased with a greater number of exposures (through vaccination, infection or exposure) (Figure 1B). Subjects with a longer interval time between last exposure and saliva collection had lower antibody concentrations than those with a shorter interval time (GMC: 28.4 IU/ml, 95% CI: 12.9−62.6 *vs* GMC: 301.3 IU/ml, 95% CI: 147.9−613.8; *p*< 0.0001) (Figure 1C). During follow-up, eight children who had two doses became infected (n=2) or were household contacts (n=6), showing an increase in antibody concentrations after exposure (*p*< 0.01) (Figure 1D). Number of vaccine doses (β1: 2314, 95% CI: 1093-3536; *p*< 0.001) and symptomatic exposure (β2: 1355, 95% CI: 340.4−2369; *p*< 0.01), were associated with antibody levels in a multivariable linear regression analysis (*p*< 0.0001).

**Table 1.** Characteristics of the study children.

|  | **Total subjects (n=89)** |
| --- | --- |
| **General characteristics** |  |
| Age, median (range), y | 10 (4–17) |
| Sex |  |
| Female, No. (%) | 42/89 (48) |
| Male, No. (%) | 47/89 (52) |
| **Exposure to SARS-CoV-2** |  |
| Unexposed, No. (%) | 36/89 (40) |
| Confirmed past SARS-CoV-2 infection, No. (%) | 14/89 (16) |
| Household contacts, No. (%) | 39/89 (44) |
| Exposed children with COVID-19 compatible symptoms, No. (%) | 30/53 (57) |
| **Vaccine schedules** |  |
| Unvaccinated, No. (%) | 1/89 (1) |
| **One dose** |  |
| BBIBP-CorV, No. (%) | 2/89 (2) |
| **Two doses** |  |
| BBIBP-CorV x 2, No. (%) | 46/89 (52) |
| BNT162b2 mRNA x 2, No. (%) | 5/89 (6) |
| mRNA-1273 x 2, No. (%) | 1/89 (1) |
| **Three doses** |  |
| BNT162b2 mRNA x 3, No. (%) | 17/89 (19) |
| BNT162b2 mRNA x 2 + mRNA-1273 x 1, No. (%) | 4/89 (4) |
| BBIBP-CorV x 2 + BNT162b2 mRNA x1, No. (%) | 11/89 (12) |
| BBIBP-CorV x 2 + mRNA-1273 x 1, No. (%) | 2/89 (2) |
| Time between most recent antigen exposure and sample collection, median (range), days | 77 (21–270) |

BBIBP-CorV (Sinopharm); BNT162b2 mRNA (Pfizer–BioNTech); mRNA-1273 (Moderna).

**Figure 1.**
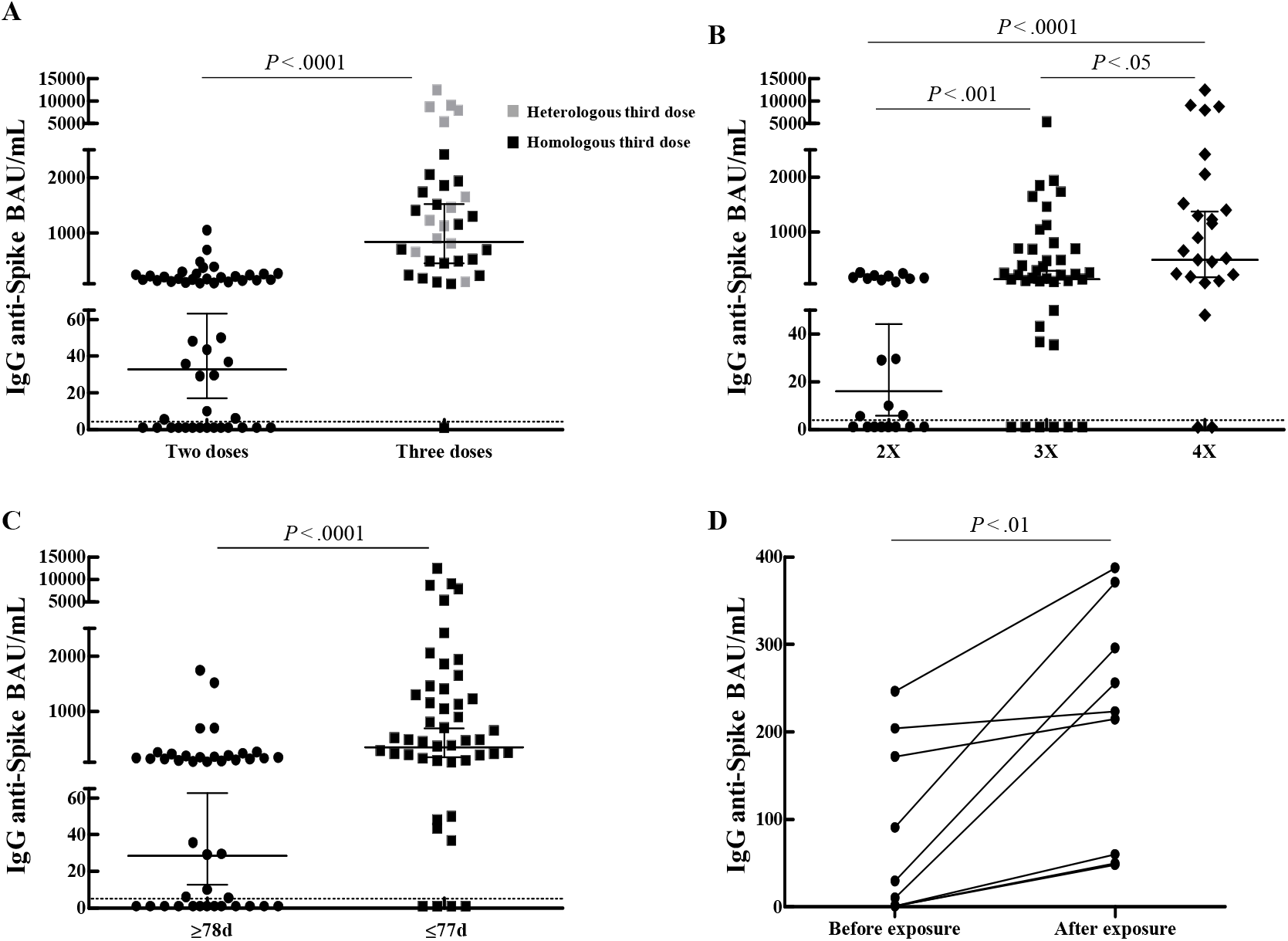
Salivary anti-SARS-CoV-2 antibodies were compared between children whit two or three vaccine doses (A), different number of exposures (B), different time between last exposure and sample collection (C), before and after exposure in children with two doses (D). Assay detection limit (dotted lines) and GMC with 95% CI are shown.

## DISCUSSION

Vaccination progressed more quickly in adults, leading to less knowledge about the immune response following vaccination in children. In this study, we observed a high salivary antibody prevalence (81%), detecting antibodies even in children whose last contact with SARS-CoV-2 antigens had been 270 days before sample collection. However, higher antibody levels were observed in children who had: three doses, a greater number of exposures and a shorter interval time between last antigens exposure and saliva collection. These results agree with those reported for the systemic humoral response in vaccinated adults, suggesting booster doses in children to maintain high antibody levels.

Limitations include lack of information on neutralizing and specific IgA antibody responses. Though, we did not have saliva-blood sample pairs for all individuals, our preliminary results in vaccinated adults show a positive correlation between specific salivary and blood antibodies. Determination of salivary antibodies against SARS-CoV-2 could be a non-invasive tool for disease surveillance, vaccination follow-up and to assist vaccination strategies against COVID-19.

## Data Availability

All data produced in the present study are available upon reasonable request to the authors

## Abbreviations

(BAU/mL): binding antibody units (BAU) per mL
(GMC): geometric mean concentrations
(95% CI): 95% confidence intervals

## Acknowledgments

The authors thank all the children enrolled in this study and their parents for their participation and collaboration. Some aspects of this work could not have been fulfilled without the generous contribution of the IIHEMA, IIE and Academia Nacional de Medicina who provide financial support to our ongoing research.

## References

1. Ketas TJ, Chaturbhuj D, Portillo VMC, et al. Antibody responses to SARS-CoV-2 mRNA vaccines are detectable in saliva. Pathog Immun. 2021;6(1):116–134. doi: 10.20411/pai.v6i1.441.

2. Healy K, Pin E, Chen P, et al. Salivary IgG to SARS-CoV-2 indicates seroconversion and correlates to serum neutralization in mRNA-vaccinated immunocompromised individuals. Med (N Y). 2022;3(2):137-153.e3. doi: 10.1016/j.medj.2022.01.001.

3. Walter EB, Talaat KR, Sabharwal C, et al. Evaluation of the BNT162b2 Covid-19 vaccine in children 5 to 11 years of age. N Engl J Med. 2022;386(1):35–46. doi: 10.1056/NEJMoa2116298.

4. Frenck RW Jr, Klein NP, Kitchin N, et al. Safety, immunogenicity, and efficacy of the BNT162b2 Covid-19 vaccine in adolescents. N Engl J Med. 2021;385(3):239–250. doi: 10.1056/NEJMoa2107456.

5. Xia S, Zhang Y, Wang Y, et al. Safety and immunogenicity of an inactivated COVID-19 vaccine, BBIBP-CorV, in people younger than 18 years: a randomised, double-blind, controlled, phase 1/2 trial. Lancet Infect Dis. 2022;22(2):196–208. doi: 10.1016/S1473-3099(21)00462-X.

6. Badano MN, Sabbione F, Keitelman I, et al. Humoral response to the BBIBP-CorV vaccine over time in healthcare workers with or without exposure to SARS-CoV-2. Mol Immunol. 2022;143:94–99. doi: 10.1016/j.molimm.2022.01.009.

