## Supplemental Information for "Prevalence of salivary anti-SARS-CoV-2 IgG antibodies in vaccinated children"

**SUPPLEMENTAL METHODS**

**Study design, participants and sample collection**

In Argentina, vaccination of children began in October 2021, with the two-dose schedule of the BBIBP-CorV (Sinopharm) vaccine for children over 3 years and the BNT162b2 mRNA (Pfizer–BioNTech) or mRNA-1273 (Moderna) vaccines for children over 12 years. Due Omicron waves, Argentina authorized third doses with mRNA vaccines for adolescents (12-17 years) and its pediatrics formulations for children between 5-11 years, in February and May 2022, respectively.

In March 2022, we began a longitudinal observational cohort study to analyze prevalence of specific salivary antibodies in children younger than 18 years. For saliva sampling, individuals spat their first saliva of the day into a tube, without drinking, brushing teeth or eating, before collection. Saliva samples were centrifuged at 17,000 xg for 10 min (4 °C) and the supernatant was stored at -20°C until used. For those children who became infected or were household contacts during follow-up, a second saliva sample was collected 21 to 30 days after exposure.

**SARS-CoV-2 antibody ELISA**

We have previously established the conditions for the determination of salivary antibodies, using saliva samples from vaccinated adults in follow-up, exposed or not to SARS-CoV-2. As pre-pandemic samples were unavailable, negative controls comprised PCR-negative saliva samples from early stages of the pandemic obtained before vaccination. Briefly, salivary SARS-CoV-2 IgG antibodies were determined by ELISA (COVIDAR-IgG) following the manufacturer's instructions without performing the first sample dilution. The plates of the assay are coated with a purified mixture of the spike protein and the receptor binding domain (RBD) of the SARS-CoV-2 (B.1 variant). Antibody concentrations in binding antibody units (BAU) per mL (BAU/mL) were obtained interpolating the OD 450 nm values of samples into a calibration curve constructed with the provided standard (400 BAU/mL).

**Statistical analysis**

A multivariable linear regression model was performed with age, sex, symptomatic exposure, number of vaccine doses, number of exposures to SARS-CoV-2 antigens, time between most recent antigen exposure and sample collection as independent variables and salivary antibody levels as dependent variable. The linear regression coefficients (β) with 95% confidence intervals (95% CI) were calculated.
